# Modelling the effect of COVID-19 mass vaccination on acute admissions in a major English healthcare system

**DOI:** 10.1101/2021.10.10.21264821

**Authors:** RD Booton, AL Powell, KME Turner, RM Wood

## Abstract

**Background:** Managing high levels of severe COVID-19 in the acute setting can impact upon the quality of care provided to both affected patients and those requiring other hospital services. Mass vaccination has offered a route to reduce societal restrictions while protecting hospitals from being overwhelmed. Yet, early in the mass vaccination effort, the possible effect on future bed pressures remained subject to considerable uncertainty. This paper provides an account of how, in one healthcare system, operational decision-making and bed planning was supported through modelling the effect of a range of vaccination scenarios on future COVID-19 admissions.

**Methods:** An epidemiological model of the Susceptible-Exposed-Infectious-Recovered (SEIR) type was fitted to local data for the one-million resident healthcare system located in South West England. Model parameters and vaccination scenarios were calibrated through a system-wide multi-disciplinary working group, comprising public health intelligence specialists, healthcare planners, epidemiologists, and academics. From 4 March 2021 (the time of the study), scenarios assumed incremental relaxations to societal restrictions according to the envisaged UK Government timeline, with all restrictions to be removed by 21 June 2021.

**Results:** Achieving 95% vaccine uptake in adults by 31 July 2021 would not avert a third wave in autumn 2021 but would produce a median peak bed requirement approximately 6% (IQR: 1% to 24%) of that experienced during the second wave (January 2021). A two-month delay in vaccine rollout would lead to significantly higher peak bed occupancy, at 66% (11% to 146%) of that of the second wave. If only 75% uptake was achieved (the amount typically associated with vaccination campaigns) then the second wave peak for acute and intensive care beds would be exceeded by 4% and 19% respectively, an amount which would seriously pressure hospital capacity.

**Conclusion:** Modelling provided support to senior managers in setting the number of acute and intensive care beds to make available for COVID-19 patients, as well as highlighting the importance of public health in promoting high vaccine uptake among the population. Forecast accuracy has since been supported by actual data collected following the analysis, with observed peak bed occupancy falling comfortably within the inter-quartile range of modelled projections.

## Introduction

At the time of the study, in March 2021, mass vaccination of the population was widely considered to be essential in exiting societal restrictions imposed during the COVID-19 pandemic, allowing a return to normal life and reopening of the travel and leisure industries and other sectors of the economy [1,2]. For healthcare systems, the reduced risk of transmission and infection through vaccination would mean lower emergency COVID-19 demand. This would be expected to reduce acute hospital pressure and enable progress towards pre-pandemic levels of elective treatment, following a period of time in which waiting times had increased significantly [3].

To facilitate effective capacity planning, for both emergency and elective care, hospitals needed to know the scale of continuing pressures they may expect from COVID-19 [4,5]. While the majority of the published literature had, at the time of the study, concerned national-level projections, there was very little evidence of COVID-19 modelling at the local level necessary to effectively guide planning within individual healthcare systems [6,7]. This was important since local intricacies, such as those relating to demographics, rurality and previously-acquired immunity, prohibit the useful abstraction of any national (or regional) level modelling. The aim of this paper is to provide an account of how modelling has been used within one healthcare system to project COVID-19 bed demand according to a range of vaccination scenarios considered plausible at an early stage of the mass vaccination effort.

The setting was the one-million resident population Bristol, North Somerset and South Gloucestershire (BNSSG) healthcare system in South West England. At the time of the study, 4 March 2021, 32% of the population had received at least one dose of either the Pfizer or AstraZeneca vaccine. With a national roadmap in place to release all societal restrictions by 21 June 2021 [8], management required an estimation of future acute COVID-19 pressures according to the envisaged vaccination rollout and an understanding of the effect of potential delays.

During the pandemic, the BNSSG system had made use of an established epidemiological modelling technique in predicting future bed requirements [9]. Projections from this model had featured within weekly updates to the BNSSG major incident Bronze Command Centre that had been stood up to address operational pressures brought about by the pandemic (Silver and Gold Command, overseeing tactical and strategic response respectively, would also receive outputs by exception). Producing the projections had been the responsibility of a system-wide multi-disciplinary working group comprising public health intelligence specialists, healthcare planners, epidemiologists, and academics. The working group convened on a weekly basis to define the scenarios to be modelled through an assessment of the latest data, including testing and public mobility data, observed bed occupancy, and any local or national developments regarding societal restrictions [10].

The remainder of this paper is structured as follows. The *Methods* section contains a description of the modelling approach, including the necessary adaptations made to incorporate the effect of vaccination, and a specification of the range of scenarios considered. The *Results* section presents the modelled COVID-19 bed occupancy projections for the 12-month period from April 2021. Finally, in the *Discussion* section, limitations and practical implications are addressed, in addition to an assessment of the modelled projections versus the actual bed occupancy recorded to October 2021 (the time of writing).

## Methods

### The model

Projections have been obtained through a Susceptible-Exposed-Infectious-Recovered (SEIR) type model, the kind of which has been routinely used through the COVID-19 pandemic as well as for outbreaks of other infectious disease [6,7]. Essentially, such models track over time the number of individuals within the considered population that are at various states of disease, such as susceptible to infection, exposed (infected but note yet infectious), infectious (able to transmit disease to others), and recovered (immune to future infection). The actual model used in this study is a more sophisticated variant of the standard SEIR model, capturing symptomatic and asymptomatic infection, acute and intensive care admission, and patient death (Figure 1). Full technical details of the model are available at [9].

**Figure 1.**
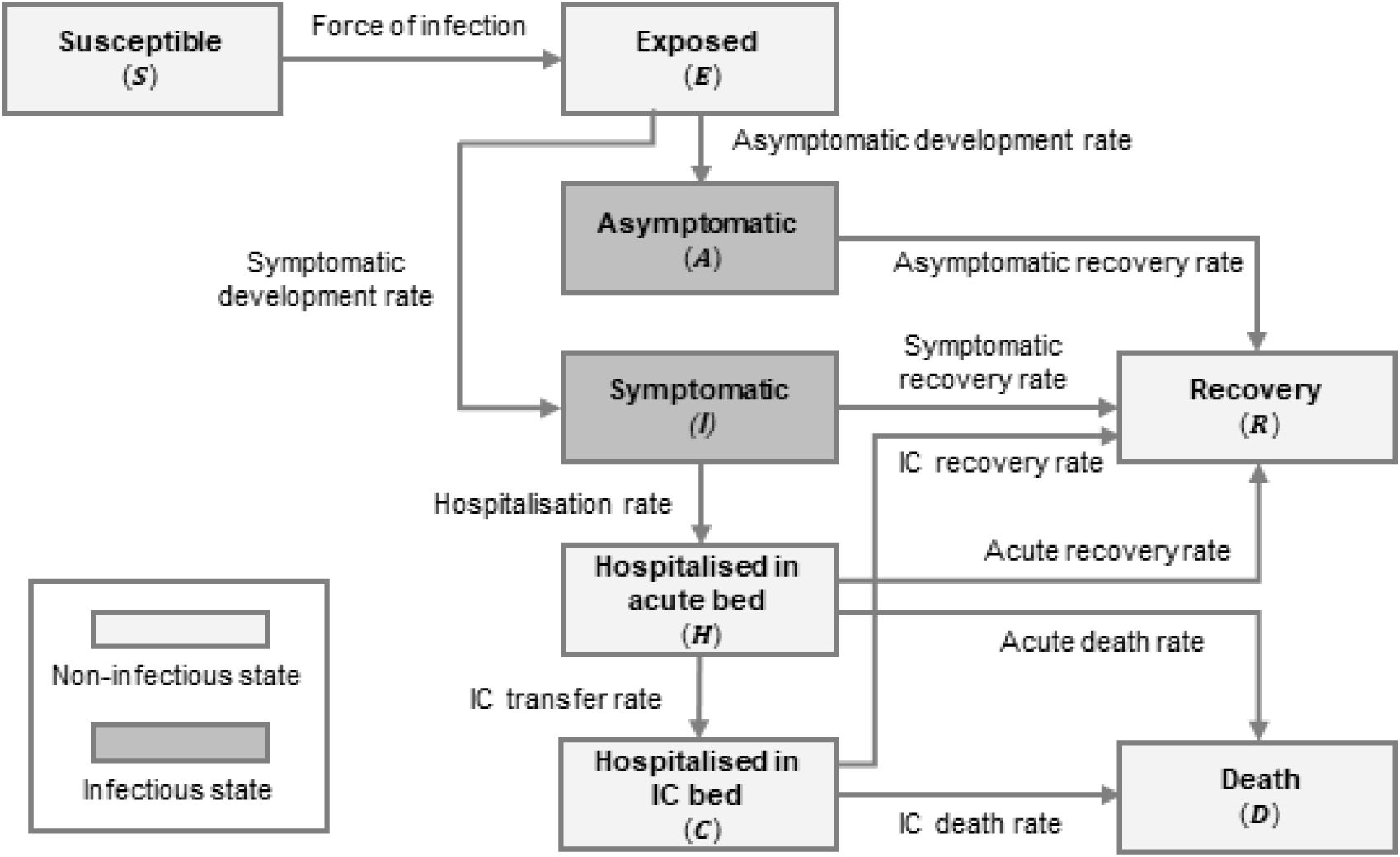
Schematic of the S-E-I-A-R-H-C-D compartmental model used within this study to project acute and intensive care bed occupancy. The model tracks the number of individuals within each ‘compartment’ over time, subject to the assumptions of each considered scenario. Note that IC refers to intensive care.

This model was adapted in January 2021 to account for the effect of vaccination, which commenced within the BNSSG system in December 2020. For calibrating the model, vaccination was assumed to reduce the likelihood of hospital admission by 95% [11] and lower transmission by 67% [12]. The model was also adapted to account for the emergence of a new variant, which had recently become dominant in the BNSSG system. The B.1.1.7 variant, also known as the Kent variant and later the *alpha* variant, was responsible for 68% of local COVID-19 cases at the end of December 2020, compared with just 12% at the start of the month. Based on information available at the time of the study, it was assumed that this new variant would increase transmissibility by 10-70% [13].

Following the same approach to calibration as for the original model [9,10], the various other social, epidemiological and clinical parameters were estimated in one of two ways. Where there was sufficiently reliably information, they were quantified based on literature findings (e.g. social contacts [14]) or local data (e.g. hospital length of stay). Where no such evidence existed, they were estimated by maximum likelihood in fitting the model to the historical bed occupancy data. In this regard, point estimates were obtained from a supplied min-max range for the given parameters, such as the aforementioned 10-70% range for the increase in transmissibility due to the new variant. This ensured that no undue confidence was placed on any particular parameter estimate. Further detail on model calibration is provided within the Supplementary Material.

### Scenario analysis

The Baseline Scenario accounts for the nationally-planned incremental relaxations to societal restrictions [8] and the envisaged vaccination rollout within the BNSSG system. According to this, order of vaccination is based upon decreasing age [15] under an assumption that all adults will be offered vaccination by 31 July 2021 and that final uptake will be 95% [16]. The date this milestone is reached is extended by two months under Scenarios 1 and 2, with the former accounting for a slower vaccination rate and the latter assuming a fixed delay at the outset (i.e. in representing possible vaccine supply issues which were of concern at the time of the study [17]). Scenarios 3 and 4 assume reduced uptakes of 85% and 75% respectively (i.e. that of typical vaccination campaigns [16]) under the same timescales as the Baseline Scenario. For context, Scenario 5 is defined by no further vaccination from 4 March 2021 and Scenario 6 assumes the envisaged vaccination rollout but with an immediate end to societal restrictions on 4 March 2021. All scenarios were modelled for the 12-month period from April 2021.

## Results

Envisaged vaccine rollout and planned relaxations to societal restrictions (the Baseline Scenario) raised the prospect of a third wave emerging in autumn 2021 (Figure 2). This is produced by increased transmission within the susceptible population who have no infection- or vaccine-acquired immunity (i.e. children and the remaining 5% of adults not vaccinated). Peak bed demand would be in late November to mid December 2021 which, given the assumption of no further imposed restrictions, coincides with ‘herd immunity’ having been reached (at this point 81% of the population have either infection- or vaccine-acquired immunity).

**Figure 2.**
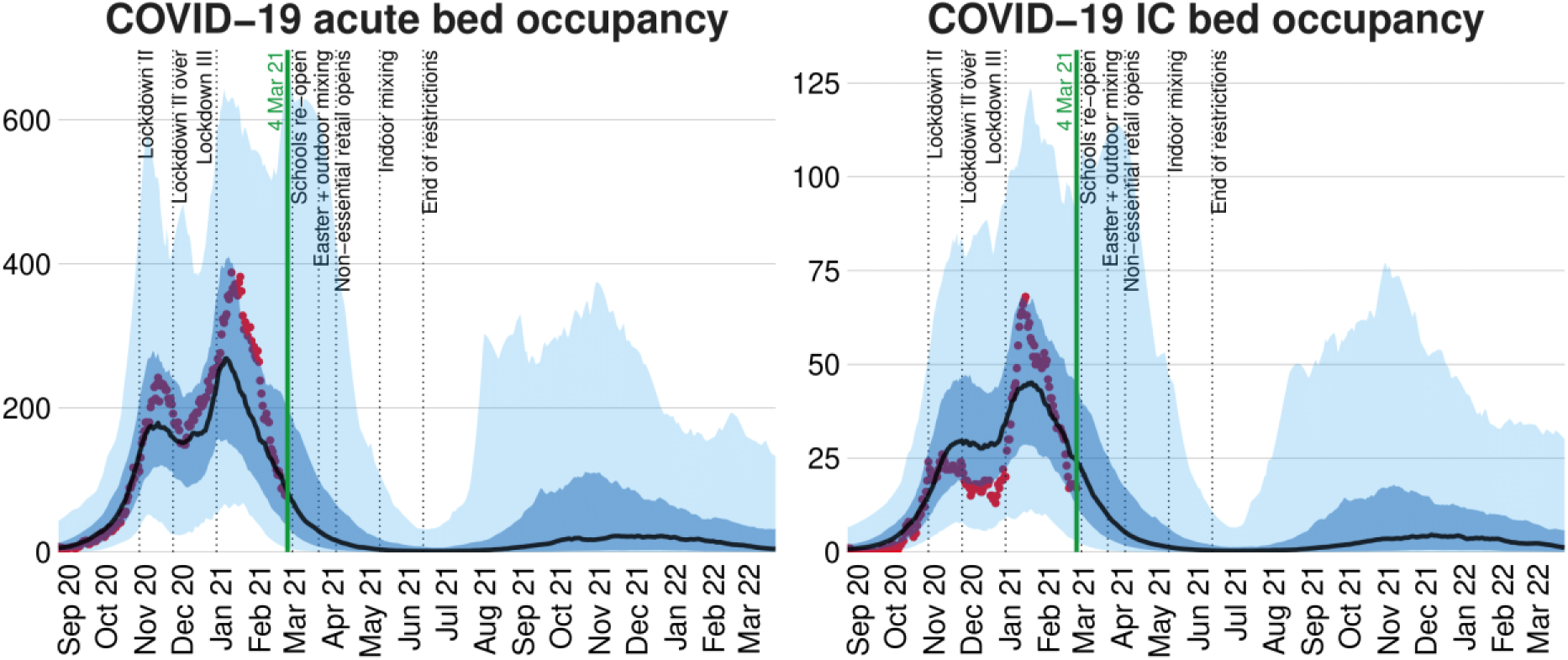
Baseline projections of COVID-19 acute and intensive care (IC) bed demand within the BNSSG system (dark and light blue areas represent the 50% and 95% credible intervals). Red dots represent historical observations of bed occupancy. The Baseline Scenario accounts for planned societal restrictions and vaccination rollout as of 4 March 2021 (when the model was run).

As would be expected – given the assumption of either or both reduced uptake or delayed rollout – all six other scenarios suggested a third wave, albeit with different timings and magnitude. The size and date of peak values are summarised in Table 1. Trajectories for Scenarios 1 to 3 – representing the most plausible alternatives to the Baseline Scenario – are shown in Figure 3. Trajectories for the remaining scenarios, all of which result in significantly higher bed demand than the second wave (peak 388 acute and 68 IC beds, observed in January 2021), are provided in Figure 4.

**Table 1.** Projections of third wave peak COVID-19 acute and intensive care (IC) bed demand for the Baseline Scenario and Scenarios 1 to 6. Peak size is presented as median and inter-quartile range.

| Scenario | Vaccination rollout | Societal restrictions | Acute bed demand peak |  | IC bed demand peak |  |
| --- | --- | --- | --- | --- | --- | --- |
|  |  |  | Size | Date | Size | Date |
| Baseline | Planned | Planned | 22 (2 - 95) | 25 Nov | 5 (1 - 14) | 14 Dec |
| Scenario 1 | Slower rollout | Planned | 34 (4 - 130) | 9 Oct | 7 (1 - 19) | 22 Oct |
| Scenario 2 | 2-month delay | Planned | 255 (43 - 578) | 8 Aug | 45 (8 - 88) | 23 Aug |
| Scenario 3 | 85% uptake | Planned | 138 (24 - 361) | 1 Nov | 29 (5 - 58) | 9 Nov |
| Scenario 4 | 75% uptake | Planned | 404 (89 - 824) | 4 Oct | 81 (23 - 144) | 15 Oct |
| Scenario 5 | Immediate end | Planned | 2250 (904 - 3624) | 6 Aug | 398 (213 - 551) | 22 Aug |
| Scenario 6 | Planned | Immediate end | 1660 (805 - 2456) | 23 Apr | 266 (93 - 404) | 24 Apr |

**Figure 3.**
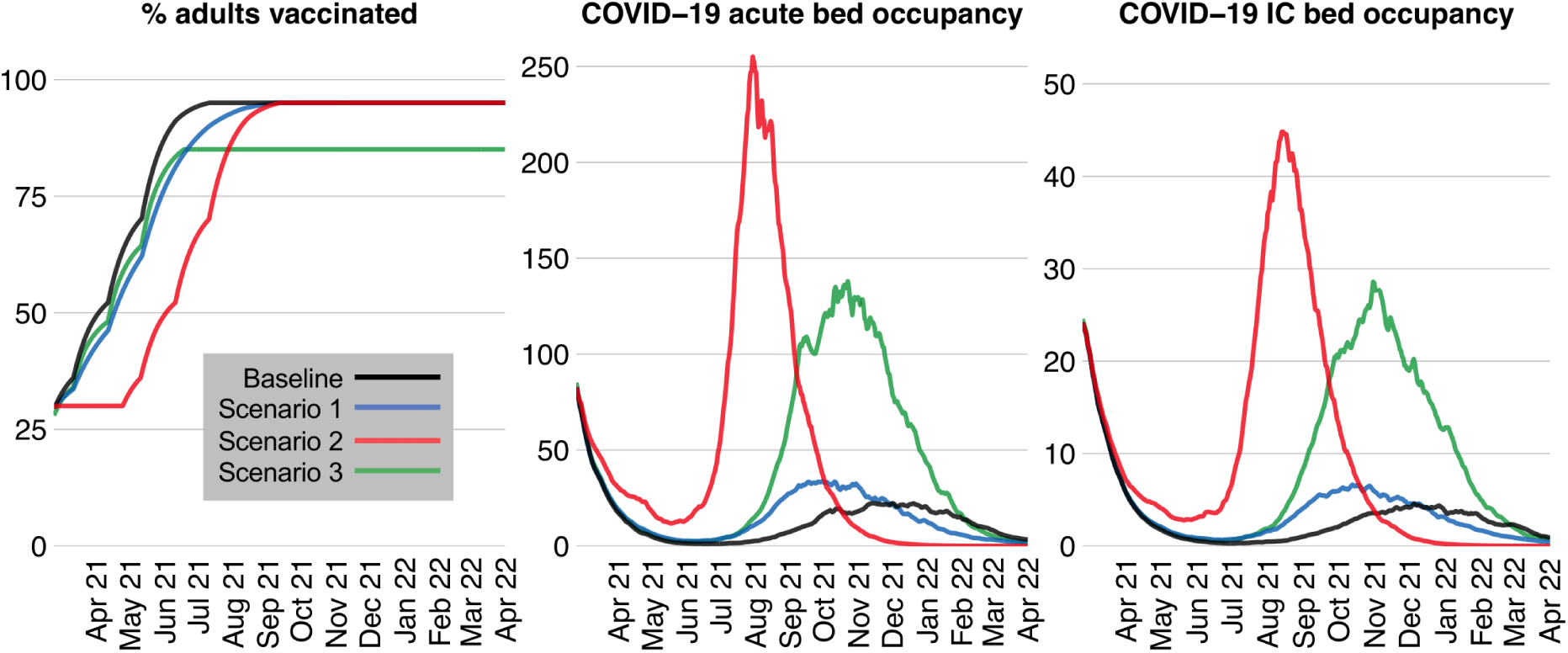
Effect of different vaccination rollouts on projected COVID-19 acute and intensive care (IC) bed demand. Results are presented for the Baseline Scenario and Scenarios 1 to 3 as modelled on 4 March 2021.

**Figure 4.**
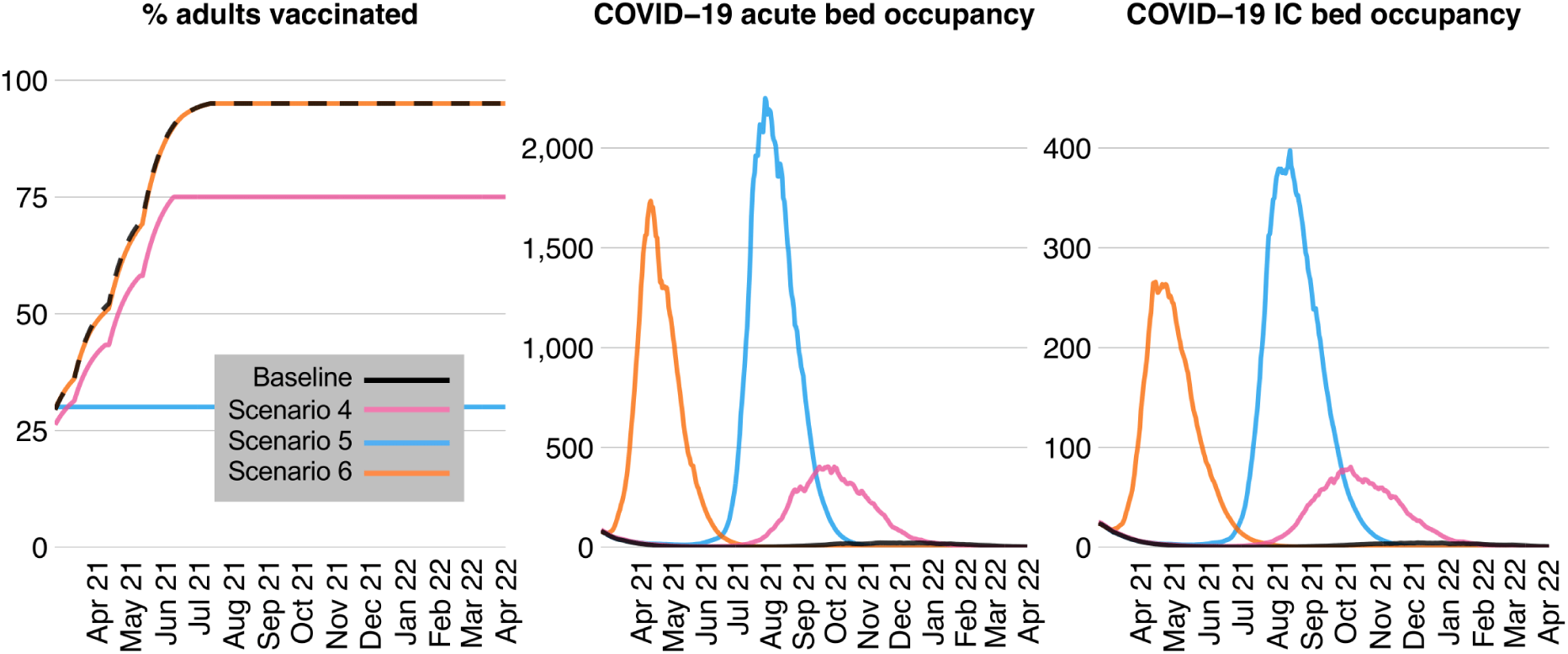
Effect of different vaccination rollouts and societal restrictions on projected COVID-19 acute and intensive care (IC) bed demand. Results are presented for the Baseline Scenario and Scenarios 4 to 6 as modelled on 4 March 2021.

## Discussion

### Statement of principal findings

Modelling supported the importance of a high vaccination uptake in averting potentially severe hospital bed pressures into the autumn and winter of 2021. Achieving 95% uptake in adults by 31 July 2021 would not avert a third wave but would ensure peak bed requirements are substantially lower than that experienced during the second wave. The second wave peak for acute and IC beds would be exceeded by 4% and 19% respectively if the typical vaccination campaign uptake of 75% was managed [16]. Such a level of bed demand would cause serious pressure on hospital capacity, and would likely require mitigation through restrictions on movement and contacts (i.e. a return to societal restrictions following full release on 21 June 2021).

Provided uptake reaches 95% by 30 September 2021, a slower vaccination rate would not lead to a substantially higher peak bed occupancy than under the Baseline Scenario but would, through faster transmission among the non-vaccinated cohort during the summer, bring forward the peak by approximately six weeks. Were vaccination delayed by two months (e.g. due to supply issues, such as those of concern at the time of the study [17]) then the rate of decrease in bed demand would reduce to the point at which all societal restrictions are released, followed by a rapid increase to a peak more comparable to that of the second wave. Results highlighted the importance of vaccinating those who may be of low hospital admission risk but who can still act as an effective transmission vector to those whose risk is greater.

### Strengths and limitations

Modelling future COVID-19 bed demand has been useful in supporting a range of operational management decisions within the BNSSG system, such as opening additional acute infection wards and procuring downstream capacity in the community (given that approximately one-fifth of BNSSG emergency acute patients require ‘step-down’ care upon discharge). Results informed such considerations within the short term, as well as others of a more strategic nature – for instance, the ability to work through the elective backlog at times of low COVID-19 bed demand, without fear of being overwhelmed. Results also informed the public health message to the BNSSG population, in stressing the importance of high vaccine uptake to avoid severe pressure on local hospitals.

Turning to limitations, it should be noted that this study did not consider any future SARS-CoV-2 variant nor assume that immunity may wane over time. While, at the time of the study, it was known that neither should be considered unlikely [18,19], there was a deficit of data required to obtain a reliable calibration. Also, due to a lack of credible data, it was not possible to model multiple vaccine doses and so it was assumed that the full benefits of vaccination derive from the first inoculation (this may have been a particular limitation given that the UK was, at the time of the study, applying a 12- week interval between first and second dose [20]).

### Interpretation within the context of the wider literature

In early March 2021, at the time of our study, there was no relevant literature regarding the impact of vaccination on hospital admissions. Acknowledging this, Moore et al published the first such investigation on 18 March 2021 [21]. In similarity with our approach, their model benefitted from age-dependent parameters (e.g. relating to probability of illness and length of stay) and was limited by equivalent assumptions on future variants and waning immunity. In line with our findings, their influential modelling, conducted at a UK national level, suggested “*that vaccination alone is insufficient to contain the outbreak*”, with a projected rise in acute admissions in the weeks and months after completing the vaccination effort. Other studies have since followed, both in the UK setting [22] and further afield [23,24].

With seven months passed since our modelling was performed, it is possible to review projections in light of what has actually happened in the BNSSG system. Given an ultimate vaccine uptake of 83%, with all adults offered vaccination by the end of July (the planned timeline), the closest scenario of those considered is Scenario 3. Under this scenario, peak acute and intensive care bed occupancies of 138 (IQR: 24-361) and 29 (IQR: 5-58) were projected, to occur in early November (Table 1). This compares favourably to the actual peaks of 105 and 21 reached within the BNSSG system, albeit a number of weeks earlier than predicted, in mid-September. At the time of writing (mid-October 2021), hospital admissions have shown positive signs of continuing their reduction, with the vaccination effort now moving onto children and third-dose boosters for adults.

### Implications for policy, practice and research

With or without vaccination, COVID-19 (or indeed another infectious disease) may pose a threat to the effective running of healthcare services, with bed modelling serving as a valuable asset to hospital managers. This paper describes an approach used to obtain such projections at a local level. For other systems looking to replicate the modelling performed here, the open-source code is available at [9], noting that other freely available models also exist [25]. While research in this field has already advanced in the months since our study, with models capturing multiple doses [21-23] and extension to children [24], there remains much opportunity for further work, not least given the significant burden COVID-19 continues to place on health services and hospital management. With much of the modelling to date remaining at the national and regional level, future investigators may wish to consider how efforts may be directed at the local level in supporting individual healthcare systems with easy-to-use, locally-configurable and reusable models; recognising that much of the actual on-the-ground decision-making takes place at this level.

### Conclusions

In providing a range of actionable insights to local decision-makers, this study has demonstrated the value of modelling in projecting the effect of COVID-19 mass vaccination on hospital bed demand. Other healthcare systems interested in adopting a similar approach should ensure the conceptual and practical suitability of any considered model, and call upon a diverse range of specialist skills and experience within the working group overseeing its use.

## Data Availability

The datasets generated during and analysed during the current study are not publicly available but are available in aggregate form from the corresponding author on reasonable request.

