## supplementary material for "Modelling the effect of COVID-19 mass vaccination on acute admissions in a major English healthcare system"

**Supplementary Material: Model adaptations and calibration**

A published model of COVID-19 transmission and bed occupancy in the BNSSG system [1] was adapted to include new epidemiological and societal developments. These include vaccination (rollout by age, uptake, efficacy and 67% reduction in transmission [2]), the emergence of a new dominant UK variant (lineage B.1.1.7 with 10-70% increased transmission [3]), reduced infectivity (37-88%) and susceptibility (31-55%) in children [4,5], and social mixing assumptions in line with UK Government announcements [6]. From 4 March 2021, 'normal' social contacts (parameterised from POLYMOD [7]) reduced according to the following 2021 timeline of relaxations to societal restrictions [6]:

8 Mar 2021: Schools reopen
9 Mar 2021: Outdoor mixing
12 Apr 2021: Non-essential retail opens
17 May 2021: Indoor mixing
21 Jun 2021: End of all legal limits on societal restrictions

A schematic of the original model is provided in Figure SM.A.1 with a full description of parameters available in [1].

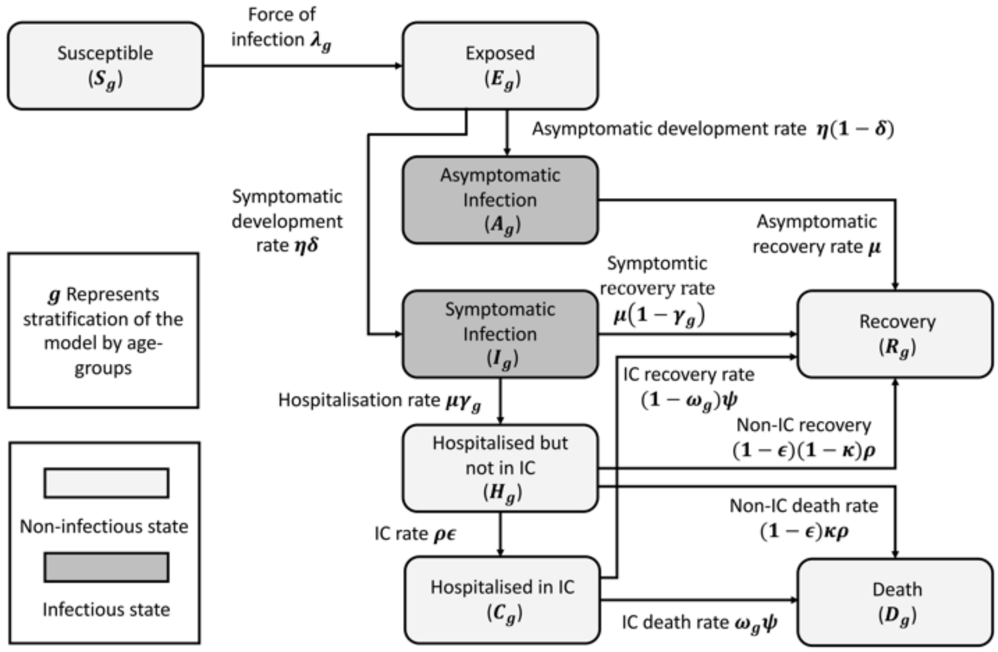


**Figure SM.1**. Model schematic from [1]**:** Compartmental flow diagram depicting stages of disease and transitions between states. Asymptomatic infection represents the number of people never showing symptoms, while symptomatic infection includes all those who show pre-symptomatic and mild symptoms to those who show more severe symptoms (pre-hospitalisation). Those who are hospitalised first occupy an acute (non-IC) bed after which they can either transfer to an IC bed, recover or die. Those in IC can either recover or die at an increased rate compared to those in acute beds.

*Effect of societal restrictions on assumed social mixing parameters*

As in [1], the POLYMOD study [7] was used to parameterise 'normal' social contacts, and to populate an age specific mixing matrix. Over time, this matrix is reduced by the introduction of various national restrictions. The restrictions included are as follows (schools closed unless otherwise indicated, resulting in an assumed 5% contact rate between <18):

To 15 Mar 2020: Normal mixing and schools open (i.e. 100% contacts)
20 Mar 2020: Social distancing encouraged and schools open (85 - 100%)
23 Mar 2020: Lockdown I (5 – 40%)
11 May 2020: Lockdown I over, distancing and mask wearing (15 – 60%)
4 Jul 2020: Further relaxations, pubs open (40 – 80%)
1 Sep 2020: Schools re-open (40 – 95% among age <18)
5 Nov 2020: Lockdown II, schools open (5 – 60%)
1 Dec 2020: Lockdown II over, tier III, schools open (10 – 80%)
24 Dec 2020: Christmas relaxation period (40 - 95%)
26 Dec 2020: Christmas mixing over (15 – 80%)
6 Jan 2021: Lockdown III (5 - 60%) – same as 5 Nov 2020
8 Mar 2021: Schools re-open (40 – 95% among age <18) = same as 1 Sep 2020
29 Mar 2021: Outdoor mixing, schools closed over Easter holidays (15 – 60%) – same as 11 May 2020
12 Apr 2021: Non-essential retail opens, schools open (15- 60%)
17 May 2021: Indoor mixing and hospitality opens, schools open (40 – 80%) – same as 4 Jul 2020
From 21 Jun 2021: End of restrictions, schools open (100%)

The % indicates the minimum and maximum range of social contexts for the optimum values to be selected through model fitting.

*Vaccination rollout by age group*

Vaccination occurs in the susceptible, exposed, asymptomatic, symptomatic and recovered compartments (but not for those who are currently in hospital in either acute or IC beds, or dead). Vaccination occurs at a rate of 5% per each age group per day, according to age dependent assumptions for start date of vaccination (based upon [8]):

8 Dec 2020: Start of vaccination for 70+ age group (including 80+)
1 Feb 2021: Start of vaccination for 60-69 age group
20 Feb 2021: Start of vaccination for 50-59 age group
20 Mar 2021: Start of vaccination for 40-49 age group
20 Apr 2021: Start of vaccination for 30-39 age group
20 May 2021: Start of vaccination for 18-29 age group

Vaccinated individuals have 95% reduced hospitalisation rate (serious illness), and 67% reduction in onwards transmission. Uptake is set at 95% – that is, when an age group reaches 95% then vaccination in that age group will cease.

*New dominant variant*

It is assumed that the aforementioned SARS-CoV-2 variant emerged on 14 Nov 2020 and linearly increased up to 23 Jan 2021 upon becoming the dominant strain. This is based on local testing data from the BNSSG system, which indicated a rise from 12% to 68% in the proportion of cases attributable to this variant within the month of December 2020. An additional increase in transmission is assumed due to the new variant of 10-70% [3]. This reflects the uncertainty in how the new variant increases transmissibility. Therefore, the new R0 under no social restrictions is 10-70% higher than the assumed 1.63 – 3.95 in the original paper [1] (Figure SM.A.2).

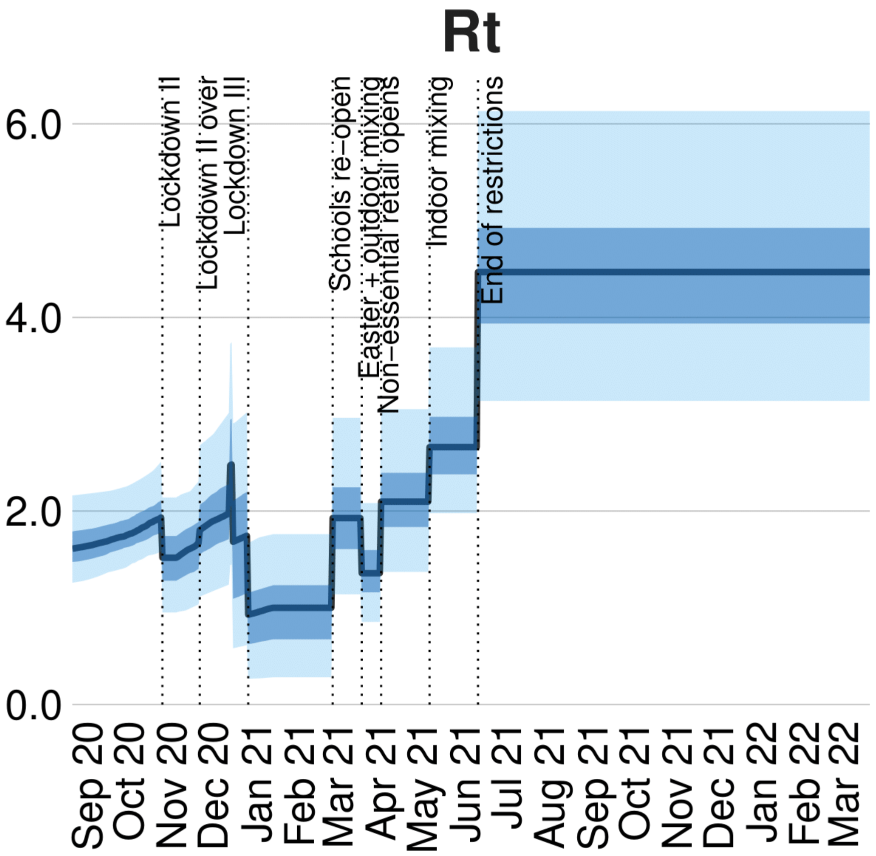


**Figure SM.2.** Estimated Rt plotted over time in accordance with the various levels of relaxations to societal restrictions as planned in [6]. R0 is equal to Rt upon full relaxation of societal restrictions from 21 June 2021.

[3] Davies NG, Abbott S, Barnard RC, Jarvis CI, Kucharski AJ, Munday JD, Pearson CA, Russell TW, Tully DC, Washburne AD, Wenseleers T. Estimated transmissibility and impact

of SARS-CoV-2 lineage B. 1.1. 7 in England. Science. 2021 Mar 3. https://doi.org/10.1126/science.abg3055.
